# Systematic investigation of gengetically determined plasma and urinary biomarkers to identify potential detection and intervention targets for vascular calcification

**DOI:** 10.64898/2026.07.31.26359444

**Authors:** Yuning Liu, Aoran Huang, Qiwei He, Bing Dong, Jinkun Cheng, Yanmeng Wu, Minjia Feng, Zhengyan Guan, Fuhong Zhu, Minhong Luo, Hui Huang

## Abstract

**Background:** Vascular calcification (VC) is prevalent among patients with atherosclerosis, hypertension, diabetes, elderly individuals, and those with chronic kidney disease. However, effective diagnostic and therapeutic strategies are lacking. Plasma and urinary biomarkers are commonly used in clinical settings to assess disease progression.

**Objective:** This study investigates causal relationships between plasma/urinary biomarkers and VC using Mendelian randomization (MR), aiming to identify potential targets for VC intervention, and to assess whether biomarkers mediate the effects of modifiable risk factors on VC.

**Methods:** We performed two-sample bidirectional MR using large-scale genome-wide association study data (n=363,228 for biomarkers, n=28,654 for VC) to investigate the causal effects of plasma/urinary biomarkers on VC. Then, a literature review was performed to identify common VC risk factors, followed by univariate MR to select risk factors associated with both VC and biomarkers. Finally, mediation analysis was conducted to explored whether biomarkers mediate the effects of modifiable risk factors on VC.

**Results:** MR analysis identified 7 plasma biomarkers linked to VC, 5 of which were positively correlated. A literature review revealed 856 VC risk factors, with 28 identified through univariate MR analysis, 11 of which correlated with identified biomarkers. Mediation analysis showed that 5 biomarkers (TRIG, GGT, CA, BILD, SHBG) partially mediated the effects of 4 modifiable risk factors on VC.

**Conclusion:** This study identifies several clinically common used biomarkers for diagnosing and treating VC, suggesting that modulating these biomarkers through lifestyle changes, such as controlling smoking, intake of milk, blood pressure and diabetes, may slow VC progression in patients.

**Lay Summary:** *What is already known about this subject?:*

1. Vascular calcification (VC) is prevalent among patients with atherosclerosis, hypertension, diabetes, elderly individuals, and those with chronic kidney disease.
2. Plasma and urine biomarkers are the most commonly used indicators in clinical practice for assessing disease progression.
3. However, the potential causal relationship between plasma and urine biomarkers and VC remains unclear.

*What are the new findings?:*

1. 7 clinically used plasma and urine biomarkers are causally linked with VC, while the occurrence of VC affects the expression of Alkaline Phosphatase.
2. A comprehensive literature review revealed 856 potential risk factors for VC. Univariate MR analysis identified 28 of these risk factors as causally linked to VC, with 11 also associated with specific biomarkers
3. Mediation analysis revealed that 5 biomarkers (TRIG, GGT, CA, BILD, and SHBG) partially mediated the effects of 4 risk factors on VC.

*How might it impact on clinical practice in the foreseeable future?:* Our findings suggest that patients may modulate the expression of 5 biomarkers (TRIG, GGT, CA, BILD, and SHBG) by controlling certain risk factors (intake of milk, smoking, diabetes, and systolic blood pressure) in daily life, thereby slowing the progression of VC.

## 1. Introduction

Vascular calcification (VC), characterized by the ectopic deposition of calcium-phosphate crystals within vascular walls, is commonly observed in individuals with chronic kidney disease (CKD), diabetes, hypertension, atherosclerosis, and the elderly^1, 2^. VC stands out as a major predictor of cardiovascular risk and plays a key role in driving disease progression in these patients^3^. VC is an active, regulated, and potentially reversible pathological process resembling bone or cartilage development^4^. The presence of calcified vessels leads to functional abnormalities, increased arterial stiffness, and reduced compliance, resulting in heart failure, left ventricular hypertrophy and myocardial ischemia. Furthermore, VC serves as a significant independent risk factor for thrombus formation and plaque rupture, playing a crucial role in the morbidity and mortality associated with cardiovascular diseases^5^. Unfortunately, effective treatment options for vascular calcification remain limited; therefore, early identification and monitoring are crucial for preventing or reducing the incidence of VC, promoting better health outcomes for patients.

Metabolites play a pivotal role as direct participants and end products of various cellular pathways, serving as crucial biomarkers that link genetic and environmental influences to disease outcomes, thereby accurately reflecting an individual’s health status^6^. Among these, plasma and urinary biomarkers are vital in the clinical practice, as their expression changes provide key insights into the complex mechanisms underlying disease disorders^7^. These biomarkers are not only the most commonly used indicators for assessing renal function but are also easily obtainable clinical specimens, characterized by ease of detection, high stability, and reproducibility. Furthermore, since plasma and urinary biomarkers are significantly affected by lifestyle factors like diet, medication, smoking, and alcohol intake, they may act as intermediaries through which various risk factors exert their detrimental effects. Understanding how to modulate these biomarkers presents promising avenues for intervention to mitigate VC in patients.

Despite numerous studies linking plasma and urinary biomarkers to VC^8, 9^, most of these investigations are observational, which presents several limitations. These studies often feature a limited number of biomarkers, single-source biological samples, and small sample sizes. Additionally, they tend to focus solely on the biomarkers themselves, without considering broader contextual factors. Furthermore, the presence of confounding factors and the potential for reverse causation bias inherent in observational research can distort results, making causal inferences unreliable and hindering the understanding of biomarkers’ causal roles in VC. In contrast, Mendelian randomization (MR) employs genetic variants that are strongly linked to exposure factors as instrumental variables (IVs), thus assessing causal relationships between exposures and outcomes. This method capitalizes on the random distribution of alleles during meiosis and the fixation of germline genetic variation at conception. Consequently, MR effectively addresses the limitations of observational studies^10, 11^. To our knowledge, comprehensive MR analyses of widely used plasma and urinary biomarkers to evaluate their causal relationship with VC, including elucidating their mediation effects on modifiable risk factors remain scarce. Research predicting and intervening on VC based on these biomarkers is also limited.

In this study, we aim to utilize MR analysis to achieve several objectives: (1) to examine the causal connection between genetic predictors of commonly used plasma and urinary biomarkers and genetic susceptibility to VC; (2) to identify metabolic intermediary biomarkers associated with VC and the risk factors linked to it; and (3) to evaluate the effect of these metabolic intermediary biomarkers on VC under the influence of modifiable risk factors. Through various MR analyses, this research seeks to elucidate the influence of plasma and urinary biomarkers on VC, expanding the diagnostic significance of commonly measured indicators in patients and identifying modifiable risk factors that could reduce VC incidence, ultimately providing insights for early prediction and intervention.

## 2. Methods

### 2.1 Research design

Figure 1 presents the overall study design. Initially, we performed bidirectional MR analysis using quantitative trait locus data for 35 biomarkers from plasma and urine, sourced from the UK Biobank, in conjunction with a meta-analysis from 16 GWAS studies on VC. This approach aimed to investigate the causal associations between plasma and urinary biomarkers and VC. Subsequently, we employed univariable MR analysis to systematically identify modifiable risk factors associated with both VC and relevant biomarkers. Finally, we clarified the metabolic intermediate biomarkers linking these modifiable risk factors to VC using multivariable MR analysis and mediation analysis.

**Figure 1.**
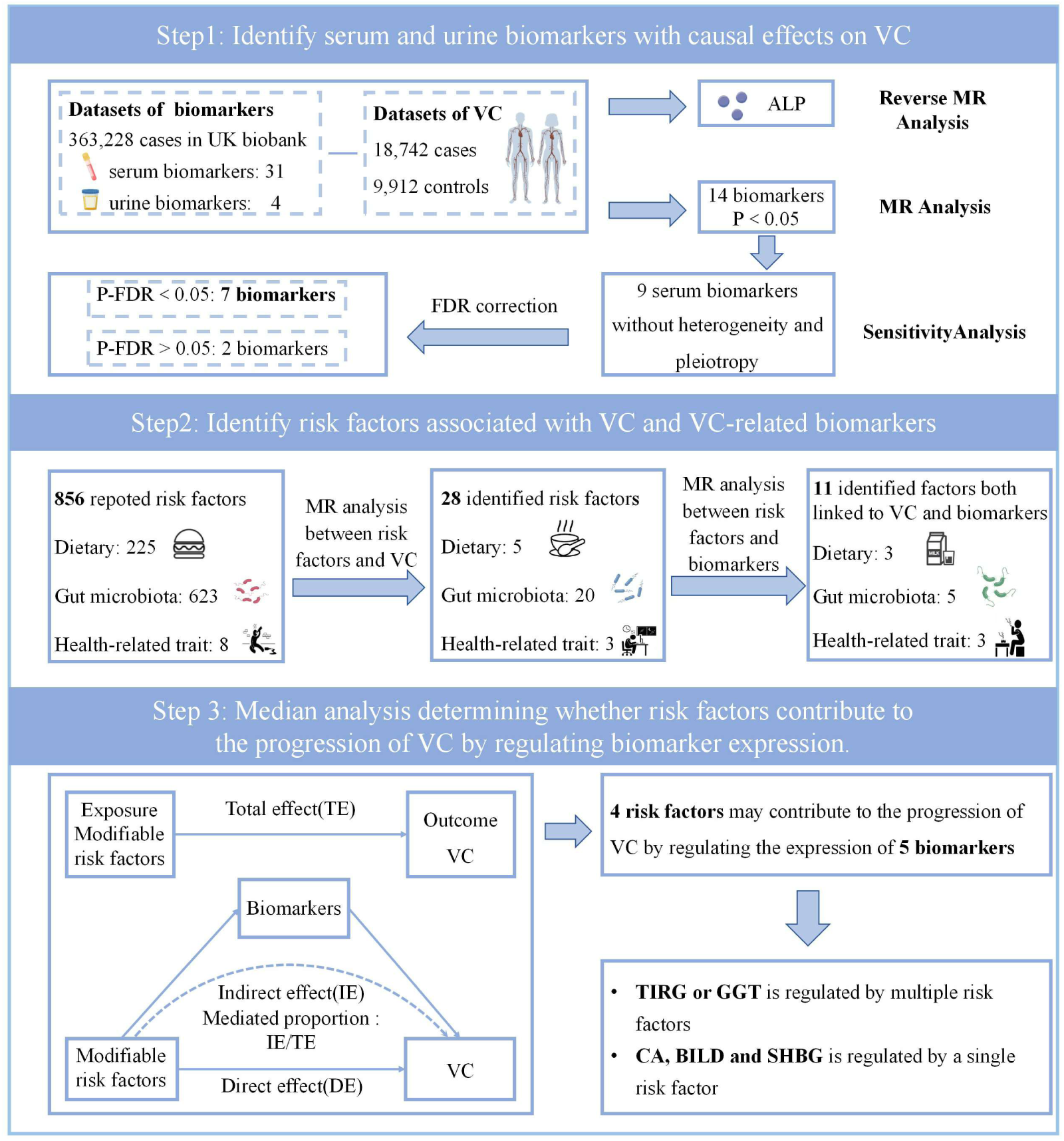
Flowchart of the study design. Step 1 provides a detailed overview of the bidirectional MR analysis investigating the causal relationships between 35 plasma and urine biomarkers and VC, ultimately identifying 7 biomarkers with causal effects on VC and 1 biomarker with a reverse causal relationship. Step 2 summarizes the process of screening modifiable risk factors associated with both VC and biomarkers using univariable MR analysis, ultimately identifying 11 modifiable risk factors from a total of 856 potentially modifiable risk factors. Step 3 describes the methods and results of the mediation analysis, revealing 5 biomarkers that mediate the effect of modifiable risk factors on VC.

### 2.2 GWAS statistics of plasma and urinary biomarkers

The UK Biobank is a comprehensive biomedical database established through a large-scale prospective cohort study. It has gathered a wide array of biological samples, genetic data, and detailed health information from more than 500,000 individuals, aged between 40 and 69 years^12^. A meta-analysis involving 363,228 UK Biobank samples accurately assessed the genetic associations of 35 plasma and urinary biomarkers, exploring their relationships with disease phenotypes and their potential role in risk stratification. Moreover, these biomarkers were quantified using an advanced liquid chromatography coupled with tandem mass spectrometry platform, further validating the high quality of this data resource. The summary statistics for the genetic associations of the plasma and urinary biomarkers utilized in this study were extracted from UK Biobank research, including 31 plasma biomarkers and 4 urinary biomarkers. Comprehensive details about the biomarker analysis can be found in Supplementary Table 1.

### 2.3 GWAS statistics of VC

The GWAS summary data for VC were acquired through a meta-analysis of 16 previously published VC GWAS study, which included data from 18,742 cases and 9,912 controls of European ancestry. Further information regarding the study population, genotyping, and imputation Can be seen in Supplementary Table 1. Informed consent was obtained from all participants, and ethical approval for the study was granted by the appropriate authorities^13^.

### 2.4 GWAS statistics of modifiable risk factors

Through a comprehensive literature review, we identified a total of 856 modifiable risk factors potentially associated with VC, including diet^14, 15^, gut microbiota^16, 17^ and health-related trait^18^. Additional information is presented in Supplementary Table 1. All participants gave their informed consent, and the study received approval from the appropriate ethical authorities.

### 2.5 Bidirectional MR analysis

We employed genetic instruments selected from the quantitative trait loci of biomarkers derived from the UK Biobank. Detailed criteria for the selection of instrumental variables (IVs) are outlined in the supplementary methods. After harmonizing the results data with VC, we retained a total of 6,986 IVs representing 35 plasma and urinary biomarkers, along with 7 IVs representing VC. Supplementary Tables 2 and 5 contain detailed information for all independent variables (IVs). Bidirectional MR analysis between biomarkers and VC was conducted using the TwoSampleMR package^19^, with analysis methods described in the supplementary materials. To address potential type I errors due to multiple testing across the 35 biomarkers, a rigorous false discovery rate (FDR) correction was applied, with the significance threshold set at p-FDR < 0.05.

### 2.6 Univariate MR analysis

We first conducted univariate MR analysis to identify risk factors with causal effects on VC (positive risk factors). Subsequently, we conducted univariate MR analysis to investigate how these positive risk factors regulate the 7 biomarkers. Risk factors that exhibited causal effects on both VC and the 7 biomarkers were utilized for subsequent analyses.

### 2.7 Multivariable MR analysis

We conducted multivariable MR analysis using the identified modifiable risk factors and biomarkers to assess whether the impact of these modifiable factors on VC is mediated through the biomarkers. Detailed descriptions of the analysis methods for this phase are provided in the supplementary methods. Finally, We calculated the mediation proportion by subtracting the direct effect from the total effect and then dividing by the total effect. All statistical analyses were conducted using R version 4.1.2.

## 3. Result

### 3.1 MR analysis identified 7 biomarkers with causal effects on VC

We conducted a MR analysis using plasma and urinary biomarkers as exposures and VC as the outcome, identifying 14 plasma biomarkers significantly associated with VC (P < 0.05) (the comprehensive MR results can be found in Supplementary Table 3). Among these biomarkers, 5 showed indications of heterogeneity or pleiotropic effects while the remaining 9 showed no such evidence (detailed sensitivity analysis results are available in Supplementary Table 4, and specific methods are outlined in the Supplementary Methods). Subsequently, we performed false discovery rate (FDR) correction on the 9 biomarkers and found that 7 plasma biomarkers, excluding TES and VITD, were statistically significantly associated with VC (P-FDR < 0.05). In summary, genetically predicted Lipoprotein a (LPA), Calcium (CA), Triglycerides (TRIG), Hemoglobin A1c (HBA1C), and Gamma-Glutamyl Transferase (GGT), which were positively correlated with VC. Conversely, Direct Bilirubin (BILD) and Sex Hormone-Binding Globulin (SHBG) exhibited negative correlations, while Vitamin D (VITD) and Testosterone (TES) were nominally significant for VC at P < 0.05 (Figure 2A).

**Figure 2.**
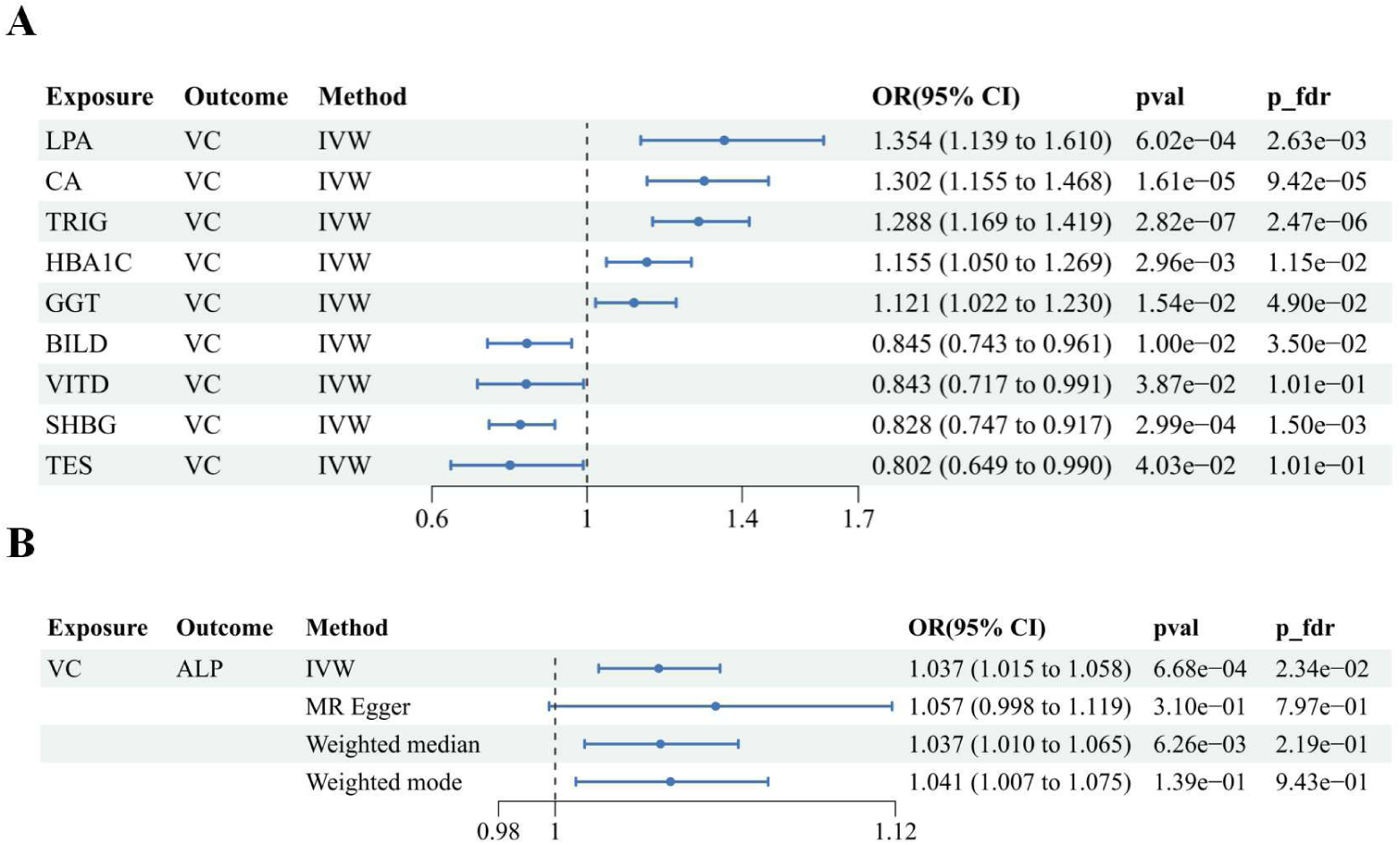
Results of the bidirectional MR analysis. (A) MR analysis with plasma and urine biomarkers as exposures and VC as the outcome, identifying causal effects of seven plasma biomarkers on VC. (B) MR analysis with VC as the exposure and plasma and urine biomarkers as outcomes, identifying the causal effect of VC on alkaline phosphatase (ALP).

Focusing solely on biomarkers as exposures and VC as the outcome may overlook potential reverse causality, which may limit the robustness of our conclusions. Therefore, we conducted a reverse MR analysis, treating VC as the exposure and plasma and urinary biomarkers as the outcomes. The results revealed a strong relationship between genetically predicted vascular calcification (VC) levels and alkaline phosphatase (ALP), which remained significant even after adjusting for the false discovery rate (FDR). In comparison, no significant associations were found with the other 34 biomarkers (detailed MR results are available in Supplementary Table 6). To verify the robustness of our results, we conducted sensitivity analyses on ALP, including leave-one-out analysis, heterogeneity tests, and horizontal pleiotropy assessments, which indicated no signs of heterogeneity or pleiotropy (see Supplementary Table 7 for details). Figure 2B illustrates the MR results for VC and ALP.

### 3.2 Univariate MR analysis identified 11 risk factors associated with both VC and biomarkers

First, we performed univariate MR analysis to assess the relationship between 856 risk factors and VC. This analysis identified 28 modifiable risk factors significantly associated with VC, including 20 gut microbiota (Figure 3A), 5 dietary factors (Figure 3B), and 3 health-related traits (Figure 3C). Detailed results are provided in Supplementary Table 9. Subsequently, we conducted univariate MR analysis to investigate the relationship between these 28 modifiable risk factors and the 7 biomarkers identified above. This analysis revealed 11 modifiable risk factors that show an association with both VC and the biomarkers, including 5 gut microbiota (Figure 3D), 3 dietary factors (Figure 3E), and 3 health-related traits (Figure 3F). Detailed results are provided in Supplementary Table 10. These findings highlight the complex interactions between modifiable risk factors, biomarkers, and VC, suggesting that these factors could serve as potential targets for therapeutic interventions in VC prevention and treatment.

**Figure 3.**
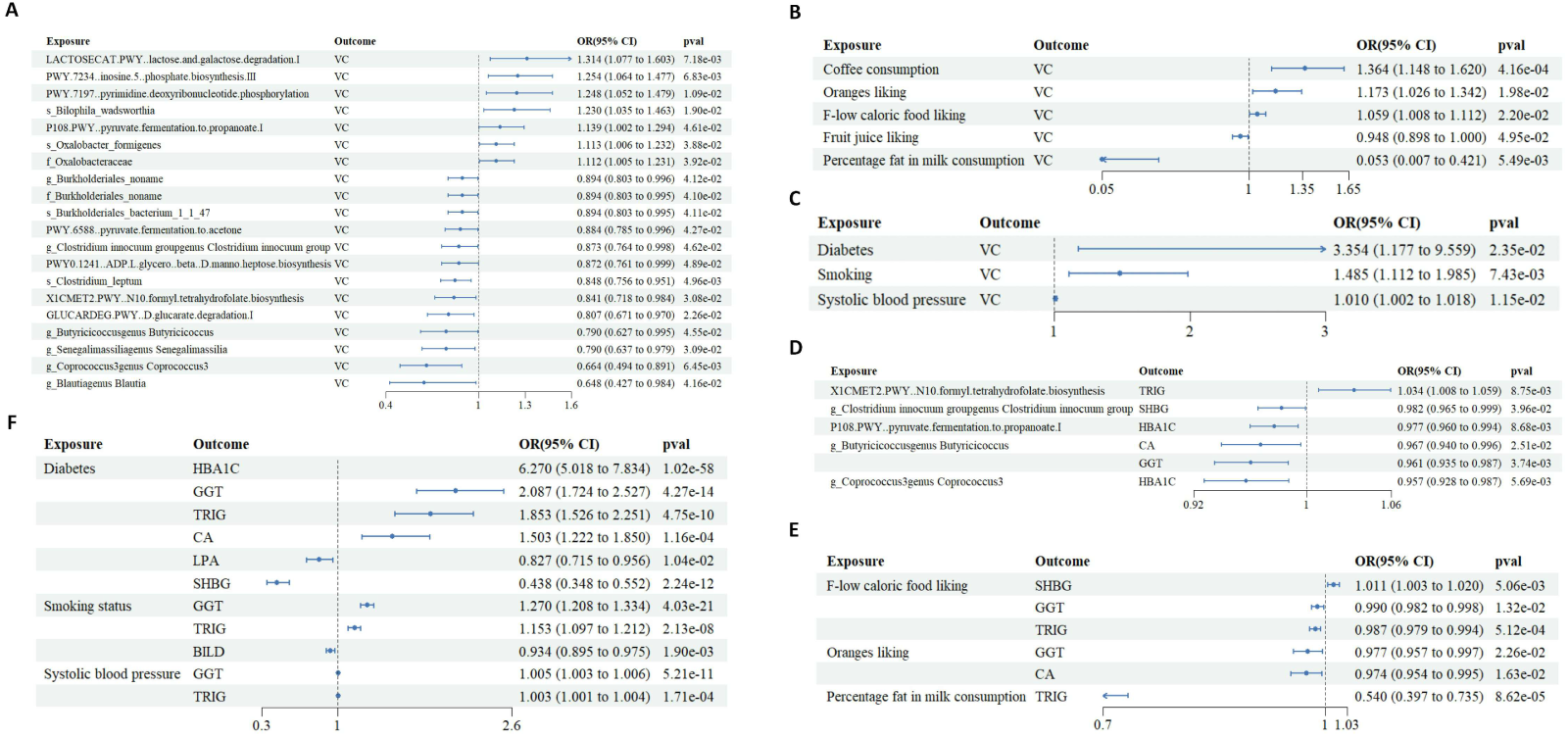
Results of the univariable MR analysis for screening modifiable risk factors. This figure presents the results of univariate MR analyses conducted with gut microbiota (A), diet (B), and health-related traits(C) as exposures and VC as the outcome, as well as the results of univariate MR analyses conducted with gut microbiota associated with VC (D), diet associated with VC (E), and health-related traits (F) associated with VC as exposures and biomarkers as the outcome.

### 3.3 Mediation analysis revealed 5 biomarkers that mediate the effect of modifiable risk factors on VC

For the sake of further investigate the complex causal relationships between risk factors, biomarkers, and VC, we performed mediation analysis involving 11 identified risk factors, 7 blood and urinary biomarkers, and VC. The results revealed that 5 biomarkers (TRIG, GGT, CA, BILD, and SHBG) mediated the effects of 4 risk factors (Percentage fat in milk consumption, Diabetes, Smoking, and Systolic blood pressure) on VC. Detailed findings are provided in Figure 4 and Supplementary Table 11. Specifically, Percentage fat in milk consumption was found to suppress VC through its effect on TRIG. Diabetes, on the other hand, promoted VC through its effects on TRIG, GGT, CA, and SHBG. Smoking also promoted VC via its impact on TRIG, GGT, and BILD, while Systolic blood pressure contributed to VC through its effects on TRIG and GGT. These findings suggest that patients could potentially reduce or slow down the progression of VC by controlling modifiable risk factors in their daily lives. By managing these factors, such as dietary fat intake, blood glucose levels, smoking habits, and blood pressure, and consequently modulating the expression of these biomarkers, it may be possible to prevent or attenuate the onset of VC.

**Figure 4.**
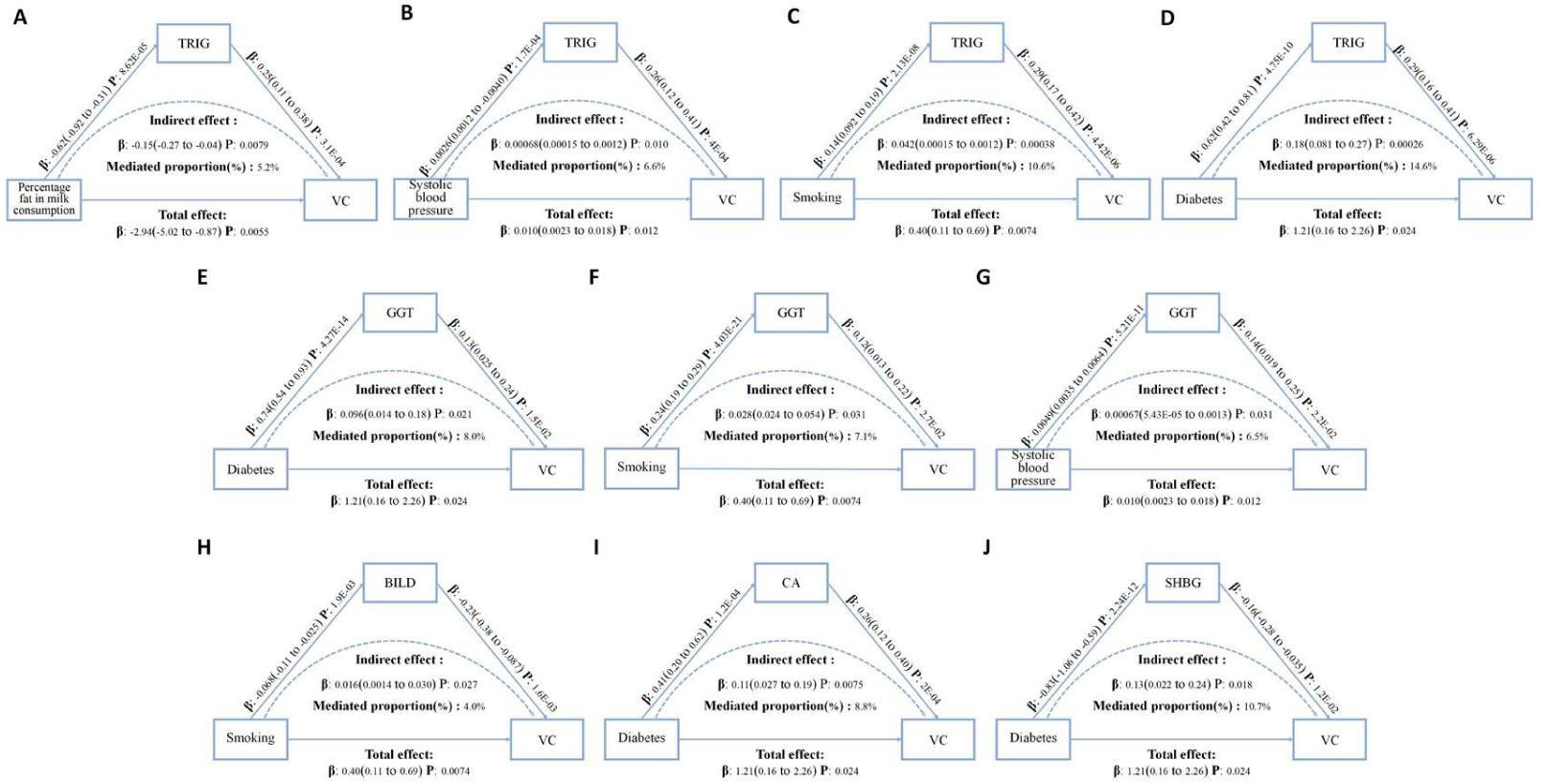
Biomarkers mediating the effect of modifiable risk factors on VC. (A) The TRIG mediates the effect of percentage fat in milk consumption on VC. (B) The TRIG mediates the effect of systolic blood pressure on VC. (C) The TRIG mediates the effect of smoking status on VC. (D) The TRIG mediates the effect of diabetes on VC. (E) The GGT mediates the effect of diabetes on VC. (F) The GGT mediates the effect of smoking status on VC. (G) The GGT mediates the effect of systolic blood pressure on VC. (H) The BILD mediates the effect of smoking status on VC. (I) The CA mediates the effect of diabetes on VC. (J) The SHBG mediates the effect of diabetes on VC.

## 4. Discussion

To explore the relationship between biomarkers and the risk of VC, we conducted a comprehensive analysis of 35 plasma and urinary biomarkers in this study. Utilizing various MR methodologies, we identified 7 plasma biomarkers significantly associated with VC. Among these, genetic prediction levels of LPA, CA, TRIG, HBA1C and GGT were positively correlated with susceptibility to VC, while BILD and VITD exhibited inverse relationships. Furthermore, our reverse MR analysis reinforced causal inferences, indicating that VC may be positively correlated with ALP levels. Given that plasma and urinary biomarkers serve as crucial early indicators of disease, their stability and accuracy can be significantly influenced by various factors. Thus, identifying and intervening in specific modifiable risk factors is of paramount importance. Mediation analysis revealed that 5 of the 7 biomarkers are modulated by 4 modifiable risk factors: Percentage fat in milk consumption, smoking status, diabetes, and systolic blood pressure. Notably, TRIG and GGT are regulated by multiple risk factors, whereas the other 3 biomarkers are influenced by single risk factors.

Our study uncovered several intriguing phenomena. TRIG may mediate the effect of four modifiable risk factors on VC, whereas GGT could serve as a mediator in the relationship between three modifiable risk factors (excluding percentage fat in milk consumption) and VC. This distinction sets TRIG and GGT apart from the other 3 biomarkers. Initially, the connection between cardiovascular disease and low-density lipoprotein cholesterol (LDL-C) was more widely recognized, in contrast to its association with triglycerides (TRIG). However, extensive epidemiological studies have shown that even after adjusting for LDL-C, TRIG concentrations remain independently linked to both the onset and progression of cardiovascular diseases^20, 21^. Current consensus suggests that elevated TRIG levels may promote VC^22^, although there is ongoing debate about whether TRIG affects VC directly, independent of other factors. Opponents argue that the connection is not robust, positing instead that it is the cholesterol content within triglyceride-rich lipoproteins (TRLs, such as Apolipoprotein B-containing lipoproteins, very low-density lipoprotein, and chylomicrons) that drives cardiovascular damage, rather than TRIG itself^23^. These TRL remnants have the potential to trigger various signaling pathways that are pro-inflammatory, pro-coagulant, and pro-apoptotic, leading to endothelial dysfunction^24^. Our findings confirm a positive causal relationship between TRIG and VC, suggesting that various modifiable risk factors can mediate this effect. This indicates that TRIG may serve as a crucial node in the mechanisms through which different factors contribute to VC, TRIG could serve as a potentially biomarker for monitoring the early onset and progression of VC, as well as the effectiveness of lifestyle interventions. Additionally, multiple cross-sectional studies have demonstrated a significant association between plasma GGT levels and various cardiovascular conditions, such as VC^25–28^, consistent with our findings. Our study clarifies the positive causal relationship of GGT in promoting VC progression. Notably, Diabetes, smoking, and elevated systolic blood pressure represent different pathogenic factors, each of which can mediate GGT’s effects and thereby promote VC. It is suggesting that, similar to TRIG, GGT could also be crucial in the pathogenesis of VC.

Interestingly, our previous clinical studies revealed a negative correlation between full-fat milk consumption and the progression of VC^29^, aligning with our current findings. Furthermore, mediation analysis involving percentage fat in milk consumption, TRIG, and VC suggests that high-fat milk intake exerts a inhibitory effect on VC, partially by mitigating the role of TRIG in promoting VC. This finding challenges the widely accepted notion that increased fat content in milk contributes to higher cardiovascular risks through fat intake. we speculate that it is because the diverse array of approximately 400 different types of fatty acids presents in full-fat milk, particularly the abundance of short-chain fatty acids, which are pivotal in preventing the progression of VC. The inhibitory effect of percentage fat in milk consumption on TRIG opens new avenues for related research, indicating that the composition and interactions of various fat components may regulate VC, potentially Offering insights that could inform the identification of novel clinical targets for the prevention and management of the condition. Exploring lifestyle interventions that involve varying fat compositions in dairy products and the consumption of full-fat milk may help reduce VC in patients during early disease stages.

In recent decades, a growing body of research has provided strong evidence supporting the beneficial effects of mildly elevated bilirubin levels against chronic conditions like hypertension, diabetes, obesity, autoimmune disorders, and metabolic syndrome^30–32^, particularly regarding cardiovascular diseases^33, 34^. A notable negative correlation exists between bilirubin concentration and VC; for instance, a Japanese study reported that an elevation of 0.058 mg/dL in plasma bilirubin concentration could reduce the probability of coronary artery calcification score ≥400 by 14%^35^. Other studies have further corroborated the inverse relationship between bilirubin concentrations and coronary artery calcification scores^36, 37^. The underlying mechanism may involve bilirubin’s ability to suppress the Multiplication of vascular smooth muscle cells and the formation of neointima^38^. This finding aligns with our conclusion that BILD has a negative causal relationship with VC.

Due to the inherent difficulties in isolating and studying the remaining bilirubin fractions, previous research often conflates total bilirubin, unconjugated bilirubin, and direct bilirubin, leading to a generalized assessment based on total bilirubin levels. Our study, leveraging the strengths of Mendelian randomization analysis, clearly elucidates the inhibitory effect of BILD on VC. Furthermore, we found that smoking can influence BILD levels, thereby promoting VC, which corroborates prior reports^39, 40^. This underscores the necessity of lifestyle interventions rather than direct modulation of biomarkers. As a crucial biomarker of liver function closely linked to cardiovascular health, BILD presents a promising target for initiating interventions to better maintain and protect overall metabolic homeostasis. Existing literature has consistently demonstrated a clear association between plasma CA levels and VC. Initially, VC was perceived as a passive biological process induced by calcium-phosphorus imbalance, which led to the deposition of hydroxyapatite crystals both in the extracellular matrix and within the layers of the arterial wall^41^. However, it is now recognized as a highly regulated and dynamic process, akin to bone formation, which includes the phenotypic transformation of vascular smooth muscle cells^42^. Epidemiological research has established a clear association between the plasma calcium-phosphate product (CaxPi) and VC in individuals with end-stage renal disease^43^. Recent prospective cohort studies also indicate that disturbances in calcium and phosphorus metabolism are key risk factors contributing to both the incidence and progression of coronary calcification in CKD patients, independent of other cardiovascular risk factors^44^. Our research confirms the positive causal relationship between CA and VC, consistent with longstanding findings in the field.

The association between SHBG and VC remains complex and often contradictory. Research indicates that sex hormones could play a role in the observed gender disparities in cardiovascular disease, potentially explaining the higher VC observed in men compared to women^45^. Several studies indicate that, after controlling for sex, age, and conventional cardiovascular risk factors, no meaningful association is found between SHBG and VC^46–48^, between SHBG levels and vascular calcification (VC). However, other investigations argue that elevated low SHBG levels in women are correlated with the advancement of coronary artery calcification, with the opposite pattern observed in men^49^. Observational studies in women further highlight a negative correlation between SHBG and both coronary and abdominal aortic calcification^50, 51^. Our MR analysis suggests a negative causal relationship between SHBG with VC, providing new insights into this long-standing debate. These findings underscore the need to consider gender differences in VC and other diseases, aligning with our review about the complex roles of sex in clinical diseases^52^. Future research should further explore the mechanisms of SHBG in VC, particularly across different genders.

ALP serves as a classic marker of osteoblastic phenotype transition and is acknowledged as a key factor in the progression of VC^53, 54^. Its activity is essential for hydroxyapatite formation during endochondral ossification, a function that parallels its role in VC, where ALP degradation of pyrophosphate, a regulator of skeletal growth and mineralization, is observed^54^. Increased ALP levels are indicative of an active osteoblastic transformation, linking external stimuli to VC pathology. Our findings clarify the causal relationship between ALP and VC, supporting the notion that VC promotes ALP production, thereby enhancing the credibility of our research.

This study offers several advantages: (1) By employing bidirectional MR, we systematically assessed the causal links between widely used plasma and urinary biomarkers and VC, minimizing reverse causation and confounding biases with a large sample size; (2) Through univariable, multivariable MR analyses, and mediation analysis, we identified various modifiable risk factors mediating the impact of biomarkers on VC, which is crucial for guiding personalized treatment, monitoring outcomes, managing chronic diseases, and promoting healthy behaviors; (3) Our findings offer valuable perspectives on the pathogenesis of VC, contributing to the advancement of predictive models based on biomarkers. However, there are limitations: (1) The study population was exclusively European, limiting the applicability of our findings to other demographic groups; (2) We did not conduct cellular or animal experiments to validate our conclusions.

## 5. Conclusion

This study identified plasma and urinary biomarkers with potential causal relationships to VC and elucidated the mediating effects of modifiable risk factors. As commonly used and cost-effective markers in clinical practice, our findings highlight new applications and clinical values for these biomarkers, offering fresh perspectives on the etiology of VC and identifying possible therapeutic targets.

## Data Availability

The data supporting the findings of this Mendelian randomization study are publicly available from the UK Biobank (https://www.ukbiobank.ac.uk) upon application. All necessary summary-level data generated during this research have been included in the Supplementary Material of this article.

## Declarations

### Author Contribution

Yuning Liu: Data collection, writing, figure preparation; Aoran Huang and Qiwei He: Data analysis, writing; Jinkun Cheng: Writing; Bing Dong: Data collection, figure preparation; Yanmeng Wu: Figure preparation; Minjia Feng and Zhengyan Guan: Writing; Minhong Luo and Fuhong Zhu: Data collection; Hui Huang: Revision, project design, funding acquisition. All authors reviewed the manuscript.

### Data Availability Statement

The GWAS data used in this study concerning plasma and urinary biomarkers, modifiable risk factors, and VC are derived from publicly available research, which can be accessed in the respective publications.

## Acknowledgements

None.

## Ethics approval and consent to participate

Not applicable

## Consent for publication

All authors have read and revised the manuscript and have approved the final version for publication.

